# COVID-19 in New York state: Effects of demographics and air quality on infection and fatality

**DOI:** 10.1101/2021.02.22.21252262

**Authors:** Sumona Mondal, Chaya Chaipitakporn, Vijay Kumar, Bridget Wangler, Supraja Gurajala, Suresh Dhaniyala, Shantanu Sur

## Abstract

The coronavirus disease 2019 (COVID-19) has had a global impact that has been unevenly distributed amongst and, even within countries. Multiple demographic and environmental factors have been associated with the risk of COVID-19 spread and fatality, including age, gender, ethnicity, poverty, and air quality among others. However, specific contributions of these factors are yet to be understood. Here, we attempted to explain the variability in infection, death, and fatality rates by understanding the contributions of a few selected factors. We compared the incidence of COVID-19 in New York State (NYS) counties during the first wave of infection and analyzed how different demographic and environmental variables associate with the variation observed across the counties. We observed that the two important COVID-19 metrics of infection rates and death rates to be well correlated, and both metrics being highest in counties located near New York City, considered one of the epicenters of the infection in the US. In contrast, disease fatality was found to be highest in a different set of counties despite registering a low infection rate. To investigate this apparent discrepancy, we divided the counties into three clusters based on COVID-19 infection, death rate, or fatality, and compared the differences in the demographic and environmental variables such as ethnicity, age, population density, poverty, temperature, and air quality in each of these clusters. Furthermore, a regression model built on this data reveals PM_2.5_ and distance from the epicenter are significant risk factors for high infection rate, while disease fatality has a strong association with age and PM_2.5_. Our results demonstrate, for the NYS, distinct contributions of old age, PM_2.5,_ ethnicity these factors to the overall COVID-19 burden and highlight the detrimental impact of poor air quality. These results could help design and direct location-specific control and mitigation strategies.

## 1 Introduction

The impact of the COVID-19 pandemic on global health and economy has exceeded well over the severity of any other communicable diseases in recent history (Baldwin and Di Mauro, 2020; Sarkodie and Owusu, 2020a). The pandemic has also stimulated and significantly accelerated global research into coronaviruses, airborne disease transmissions, and development of new vaccines. Within a short span of time, scientists have succeeded in obtaining critical information on the structure and genomic sequence of the virus pathogen SARS-CoV-2, mechanism of virus infection to host, modes of transmission, and injury to host organs induced by the virus. The research findings have accelerated the development of vaccine and established preventive measures such as the use of masks. Simultaneously, there has been a significant effort to understand the association of COVID-19 to demographic and environmental factors, to explain the geographical or seasonal variability in disease burden (Goldstein and Lee, 2020; Karmakar et al., 2021; Perone, 2021; Sorci et al., 2020).

Among the demographic variables, age, gender, ethnicity, and population density are reported to have significant impact on COVID-19. Fatality from COVID-19 was shown to significantly increase with advanced age. A study conducted on hospitalized patients in the NYC area found 84% of the total deaths occurred in people aged above 60 years (Mesas et al., 2020; Richardson et al., 2020). Moreover, males were seen to be more susceptible to suffer from COVID-19 complications and fatality (Pradhan and Olsson, 2020). Although the mechanism underlying such predisposition of age and sex is not completely understood, presence of preexisting health conditions and a lowered immunity associated with higher age are thought to be two major factors (Mesas et al., 2020; Pradhan and Olsson, 2020; Richardson et al., 2020). Chronic comorbidities such as hypertension, ischemic heart disease, diabetes, and chronic obstructive pulmonary disease (COPD) are more common in older age and poses risk for severe outcomes (Lusignan et al., 2020; Richardson et al., 2020). Studies focused on the impact of COVID-19 on the ethnic composition also revealed certain ethnicities’ vulnerabilities to the disease. In the US, a disproportionately higher number of COVID-19 infections and mortality are observed among African American and Hispanic American relative to their share of population (Martinez et al., 2020; Yancy, 2020). Socioeconomic disparities leading to increased exposure and lower access to healthcare are thought to contribute to such vulnerability. High population density is reported to increase the risk of COVID-19 spread (Arif and Sengupta, 2020; Copiello and Grillenzoni, 2020), although it is not the sole determining factor as many dense metropolitan cities in Japan, South Korea, China, and Singapore have observed a low infection rate (Lee et al., 2020; Rocklöv and Sjödin, 2020).

The association of environmental factors such as air quality and meteorological parameters to the adverse effects of COVID-19 has been investigated in multiple studies. Air pollution is of particular interest as, chronic exposure to air pollutants is linked to multiple chronic respiratory and cardiovascular diseases such as COPD, ischemic heart disease, and hypertension—diseases which are known to increase COVID-19 fatality (Feng et al., 2016b; Guan et al., 2016; Wellenius et al., 2012). Additionally, air pollution substantially increases the risk of respiratory infections including viral infections (Chauhan and Johnston, 2003; Feng et al., 2016a). Fine particulate matter in the air, especially PM_2.5_ (particulate matter with aerodynamic diameter 2.5 µm or less) have been linked to many of these pollution-mediated health effects (Brook et al., 2010; Hopke et al., 2019; Xing et al., 2016). Early reports indicate a positive association of PM_2.5_ with both COVID-19 transmission and fatality (Gupta et al., 2020; Lolli et al., 2020; Pozzer et al., 2020; Wu et al., 2020). Analysis of meteorological factors based on the data from 30 Chinese cities revealed low temperature, less diurnal temperature variation, and low humidity favors the transmission of COVID-19 infection (Liu et al., 2020). This finding was supported by a larger scale study using data from the top 20 countries with infections, and further claimed low wind-speed, surface pressure, and precipitation to increase the risk of disease spread (Sarkodie and Owusu, 2020b).

The impact of COVID-19 in infection or fatality shows a wide variation across countries or even between different regions within a country (Auger et al., 2020; Miller et al., 2020). The current body of research strongly suggests the influence of both demographic and meteorological factors underlying such differences; however, their relative contribution is not well understood. While the availability of such information will be extremely useful to develop preventive and mitigative strategies, wide variability in multiple other factors such as testing and screening strategies, healthcare infrastructure, and socio-cultural practices would play as confounding variables and hinder such analysis. Considering such limitations, in this work, we conducted our study using data from New York State (NYS). NYS offers a wide range of variation in its demographic and environmental landscapes with urban, population-dense, ethnically diverse counties near NYC to many rural, white-dominated, population-sparse counties located elsewhere. However, state-wide policies help to reduce the differences due to the confounding factors mentioned above. Using publicly available data, we grouped the counties into clusters based on COVID-19 infection, death, or fatality, and investigated their association with various demographic and environmental variables. Furthermore, regression models were built to understand the relative contribution of these risk variables on COVID-19 infection, death, and fatality.

## 2 Methods

### 2.1 Study Area, Data Source, and Variables

For this study, we acquired the infection and death count from COVID-19 data for all 62 counties in the NYS from publicly accessible information available at Syracuse.com. The data is from March 1, 2020 and May 16, 2020. The population estimates for each county were obtained from the 2018 US Census Bureau’s American Community Survey (ACS) website (https://www.census.gov/programssurveys/acs) (CDC and Team, 2020; Roser et al., 2020). Infection and mortality rates from COVID-19 were calculated by dividing the infection and death counts by the total population for the county, respectively, and expressed as number per 100,000 population. The case fatality rate (CFR) was obtained by dividing the death count by infection count and presented as number of deaths per 10,000 infected population. In addition to the total population, the ACS census database was used to collect the following information for each county:(1) Area; (2) population with age ≥ 55 years; (3) poverty levels; (4) Hispanic American population (Martinez et al., 2020); and (5) African American population (Yancy, 2020). From this information (1) population density (population/square mile), (2) proportion of population with ≥ 55 years (expressed as %), (3) proportion of Hispanic American (expressed as %), and (4) proportion of African American (expressed as %) population was calculated for each county. All factors except population density and distance from the epicenter were converted to percentages by county. The nursing home locations across the NYS counties were obtained from the Department of Health and Human Services. The data was retrieved through ArcGIS Map 10.7.1 (Monmonier and Giordano, 1998). The distances of a county from Manhattan (considered as the disease epicenter) were calculated by measuring the distance between the centroids of two locations using ArcGIS Map 10.7.1 software. The temperature and Air Quality Index (AQI) information obtained from Environmental Protection Agency (EPA) measurements available through the United States EPA website (http://www.epa.gov/ttn/airs/aqsdatamart). The monthly average PM_2.5_ estimates over the entire US were made at 0.1° X 0.1° grid resolution through a combination of satellite-derived estimates, ground based measurements, and their statistical fusion through a geographically weighted regression model (Donkelaar et al., 2019). This data was further aggregated to the geographical confinement of a county and temporally averaged for the years 2000 – 2016 (Wu et al., 2020). This county-specific, temporally averaged PM_2.5_ data was used in this study.

**Table 1.** Publicly available data sources used in this study

| Data | Source |
| --- | --- |
| Covid-19 cases & deaths | Coronavirus in NY: Cases, maps, charts, and resources<br>(https://www.syracuse.com/coronavirus-ny/) |
| Population estimates & demographics 2018 | US Census Bureau’s American Community Survey (ACS)<br>(https://www.census.gov/programs-surveys/acs) |
| Temperature & air quality index | EPA (http://www.epa.gov/ttn/airs/aqsdatamart) |
| Nursing homes locations | The Department of Health and Human Services (HHS)<br>(https://www.arcgis.com/home/item.html?id=b3813b2d3a054c378247bf32bcd8d203) |
| Satellite PM<sub>2.5</sub> estimates | Air pollution and COVID-19 mortality in the United States, Harvard University<br>(http://github.com/wxwx1993/PM_COVID) |

### 2.2 Statistical Analyses

#### 2.2.1 K-Means Clustering

The counties of NYS were classified into categories using k-means clustering technique. They were partitioned into three disjoint clusters based on their infection and mortality rate to identify any common pattern among the counties within a cluster. For the implementation of clustering algorithm, the value of k was set in advance along with the assignment of initial centroid positions for the clusters (Fahim et al., 2006). The algorithm started with the random initialization of the positions of centroids and was followed by two steps. The first step assigned each sample to its nearest centroid. The second step created a new centroid by taking the mean value of all the samples assigned to each previous centroid. The difference between the old and the new centroids were computed, and the algorithm repeated these last two steps until this difference was less than a threshold. The model used Euclidean distance for the calculation of the distance and the threshold considered was 0.0001. In the end, the centroids were fixed, did not move anymore, signifying the convergence criterion for clustering and resulted in three distinct clusters. For the fatality group, cumulative positive cases and cumulative deaths were collected from each county until 16th May 2020. The number of positive cases per 100K population and number of deaths per 10,000 positive cases were used in k-means clustering technique, considering them as two dimensions. Three distinct clusters were identified in the CFR group.

#### 2.2.2 Tests for Significance

The descriptive analysis was performed for all the parameters considered in this study. Statistical comparisons between the demographic and environmental variables in clusters were performed using Kruskal-Wallis (KW) test, a non-parametric equivalent of one-way analysis of variance (ANOVA), since the variables were non-normally distributed. Once the KW test statistic was found to be significant, multiple comparisons were conducted using Mann-Whitney U test after making the Bonferroni corrections. All analyses used 2-sided statistical tests and P < 0.10 was considered as significant.

#### 2.2.3 Autoregressive Integrated Moving Average (ARIMA) Model

An ARIMA model was used to analyze the multi-year time-series data of temperature and AQI to reduce noise and obtain better estimates. In ARIMA model, future value of a variable is predicted by a linear combination of past values and errors (Hyndman and Athanasopoulos, 2018). The model is often expressed as ARIMA (*p, d, q*), where *p, d*, and *q* represent the order of auto-regression, the degree of trend difference, and the order of moving average, respectively. The model is essentially a combination of three parts: (1) The first part is the auto-regressive model, which uses the linear combination of past values of the variable to forecast the next value and referred as an AR(p) model, an autoregressive model of order p. (2) The second part is the integrated (I), which is computed by taking the difference between the consecutive observations to make the data stationary. (3) The third part is the moving average (MA) model, referred as MA(q) and equivalent to a regression model that involves past forecast errors as predictors.

Augmented Dickey-Fuller (ADF) unit-root test was used to estimate the stationarity of each time series data that is the absence of fluctuation or periodicity with time. To build ARIMA models for temperature and AQI, data collected by EPA over a span of 5 years (2015-2019) were used. Outdoor temperature recorded hourly and AQI recorded daily were first converted into weekly data. ARIMA model requires stationary time series meaning the time series shows no fluctuation or periodicity with time. To this effect, stationarity of the data was confirmed by ADF test prior to use for model building. Three models using each of three rates were created with 95% confidence bands for each of the three clusters defined in this study. The model goodness of fit was evaluated by calculating the Akaike information criterion (AIC) for each model.

#### 2.2.4 Regression Models

Variables for the regression models were first evaluated for normality of distribution by visual inspection of histograms followed by the Shapiro-Wilks test for normality. The univariate method of outlier detection was used to eliminate outliers in the predictors. Correlation between variables was examined by constructing correlation matrices and evaluating Pearson’s correlation coefficients between the predictors. Multicollinearity between the independent variables was assessed using the variance inflation factor (VIF), which measures the inflation in the variances of the parameter estimates due to multicollinearity. An upper cut-off value of VIF for predictor variables was set as 5 to minimize the contribution of multicollinearity in our model (Chatterjee and Simonoff, 2013). A stepwise forward selection procedure was implemented to evaluate the relative contribution of predictor variables for infection, death, and fatality from COVID-19. The forward selection algorithm for stepwise regression starts with an empty model, and one variable is added at a time along with the measurement of its accuracy. This process is repeated until all variables are incorporated into the model. The goodness of the model is interpreted by the adjusted R^2^ value and the contribution of an individual variable is assessed from the order the variable was entered in the model. The residuals of the regression models were checked for model adequacy and outliers were removed when needed.

All analyses were performed using version 3.6.9 of the Python programming language.

## 3 Results

### 3.1 Distribution of COVID-19 in NYS counties

The infections and deaths from COVID-19 between March 1, 2020 and May 16, 2020 for all 62 counties in the NYS were included in the analysis. This time window roughly corresponds to the first COVID-19 wave observed in NYS. The counties were classified into three distinct clusters based on infection and death rates calculated for each of those counties using k-means clustering technique. Cluster **1** includes counties with a high infection or mortality rate, cluster **3** incorporates counties with a low rate, and cluster **2** being the intermediate between the other two (Figure 1A,1B). We observed that the cluster **1** for infection rate, where the infection ranged 2,500-4,000 per 100,000 population, consisted of 8 counties (Rockland, Westchester, Bronx, Nassau, Suffolk, Staten Island, Orange, and Queens), all located within close proximity in the downstate NY (Figure 1C). Cluster **2** was formed by 6 nearby counties, namely Ulster, Dutchess, Putnam, Manhattan, Sullivan, and Brooklyn (Figure 1C). The counties of upstate NY fell in the cluster **3** where the infection rate was <500 per 100,000 population, well below the other two clusters. The clusters for death rate showed a similar distribution across the NY states. Four counties in downstate NY, namely Bronx, Queens, Rockland, and Brooklyn were included in the cluster **1** with death rate ranging from 175 to 200 per 100,000 population (Figure 1B,1C). Of the remaining counties, 6 neighboring counties belonged to cluster **2**, and the rest of upstate counties were included in the cluster **3** with a death rate <50 per 100,000 population. Visual inspection revealed higher COVID-19 death rates are from the counties located in downstate NY (Figure 1C).

**Figure 1.**
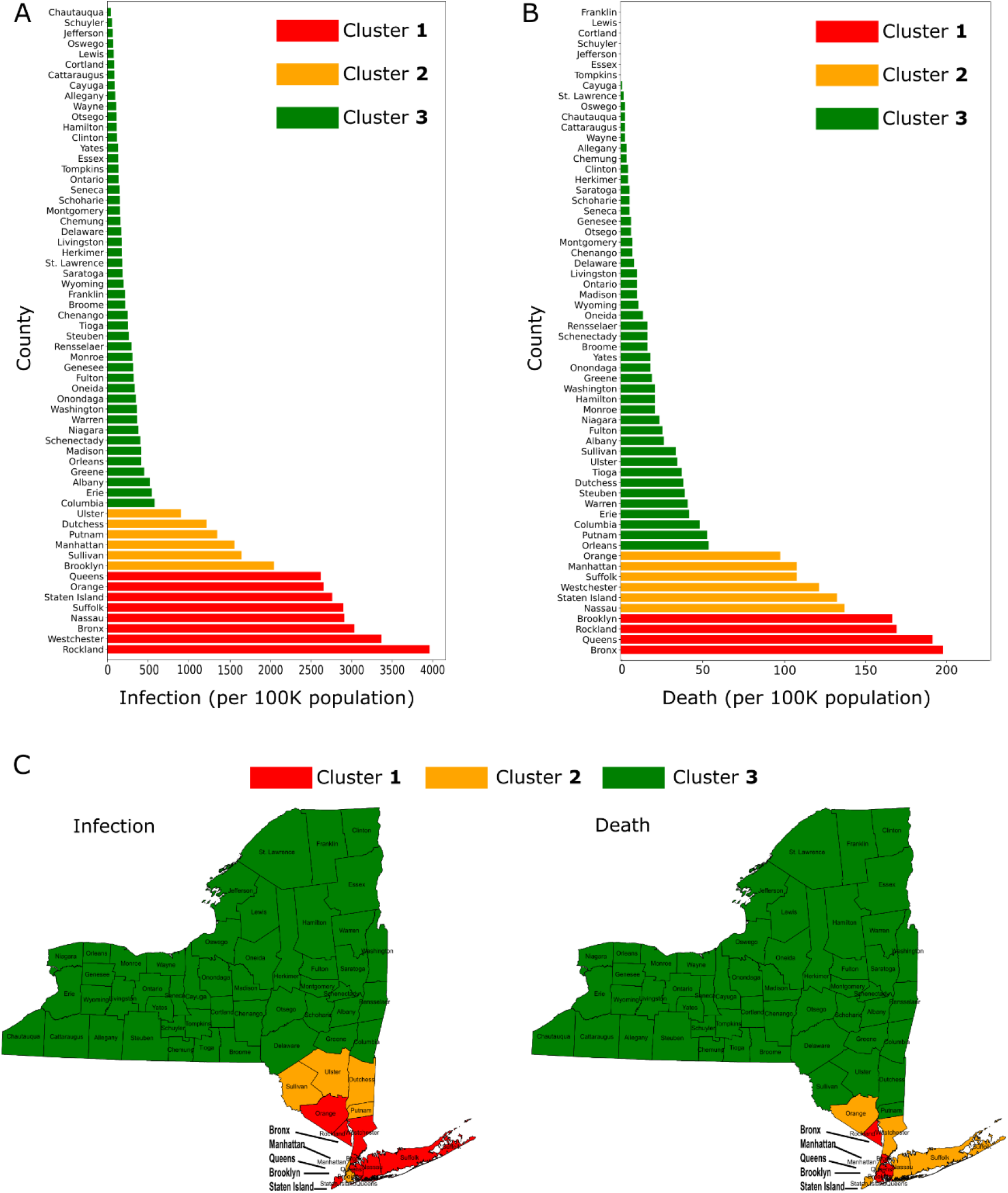
Infection and death from COVID-19 in NYS counties (data till May 16, 2020).(A-B) Infection rates (A) and death rates (B) rates of individual counties are shown; counties are further grouped into three clusters using k-means clustering technique. (C) Maps of NYS showing the locations of counties included in each of the clusters.

A similar pattern in the distribution of COVID-19 infection and death rates among NYS counties suggests an association between these two variables, which was confirmed from the scatter plot (Figure 2A) and a strong positive correlation (Pearson’s correlation; r = 0.92, P < 0.0001). The observation suggests that the number of infections in a county is a key factor influencing the number of COVID-19 deaths.

**Figure 2.**
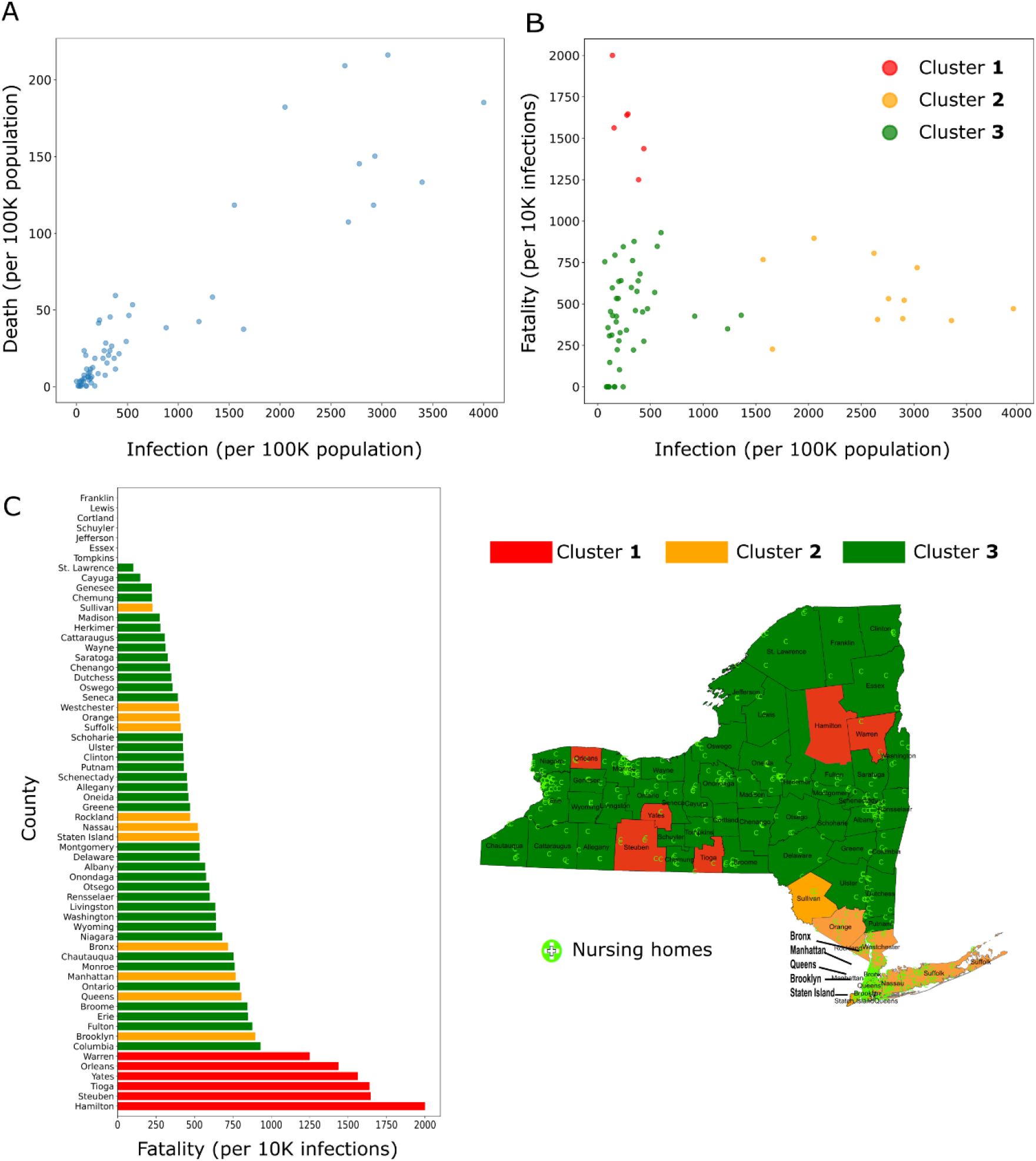
(A) Scatter plot showing the relationship between COVID-19 infection and death rates in NYS counties. (B) Fatality rate is plotted against infection rate for each county; the counties are further grouped into 3 clusters using two-dimensional k-means clustering method.(C) Distribution of counties from each cluster when sorted by fatality rate *(left)* and their location in the NYS map *(right)*. Locations of nursing homes are also depicted in the map.

Even though we observed a strong correlation between the infection and death rates, this data does not provide information about the disease fatality, defined as the proportion of deaths occurring from infections. When the fatality rate (expressed as deaths per 10,000 infections) was calculated for all counties and plotted against the infection rate, a distinct pattern of relationship between these two variables was found (Figure 2B). We observed that the counties with a high fatality rate had a relatively low infection rate while the counties with a high infection rate had a relatively low fatality rate. In accordance, when the counties were divided into three clusters on the basis of infection and fatality rate using a two-dimensional k-means clustering method, we obtained clusters with the following features: (1) High fatality and low infection rate (cluster **1**); (2) high infection and low fatality rate (cluster **2**); (3) low infection and low fatality rate (cluster **3**). Interestingly, the locations of the counties included in cluster **1** (Hamilton, Steuben, Tioga, Yates, Orleans, and Warren) were distributed across the NYS (Figure 2C). The counties in cluster **2** were all in proximity and located near NY city, although in term of fatality rate, they were interspersed with cluster **3** (Figure 2C). These results suggest that various risk factors for COVID-19 have a differential contribution on infection and fatality.

### 3.2 Impact of demographic factors on COVID-19

Multiple studies have shown the association of demographic variables with COVID-19 infection and outcome (Goldstein and Lee, 2020; Karmakar et al., 2021; Perone, 2021; Sorci et al., 2020). We selected five well-known demographic risk factors namely, population density, age (percentage of people with age above 55 yr), ethnicity (percentage of African American and Hispanic American population), and poverty (percentage with below poverty line) to study how these variables varied across NYS counties. Additionally, we considered the distance from the disease epicenter, measured as the distance of a county from Manhattan. Figure 3 shows these variables plotted against counties organized in three clusters as described in the previous section. Each variable demonstrated a characteristic pattern of distribution among the clusters. KW test followed by multiple comparison was further performed for statistical difference.

**Figure 3.**
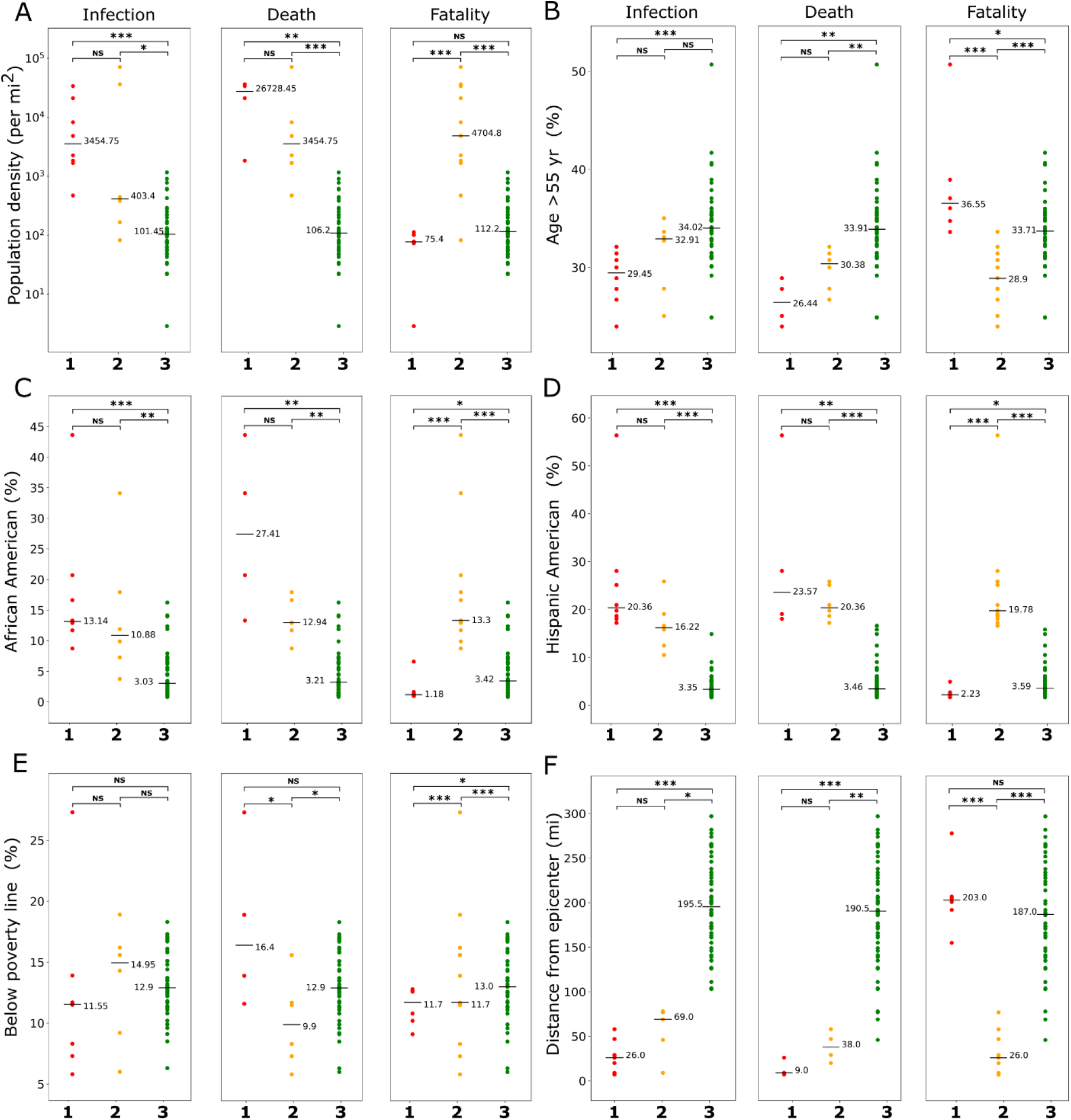
Association of demographic variables with COVID-19. For infection, death, and fatality from COVID-19, (A) population density, (B) age (> 55 yr), (C) African American population, (D) Hispanic American population, (E) population below poverty line, and (F) distance from epicenter were compared among three clusters. Horizontal bars represent medians. ***P < 0.01, **P < 0.05, *P < 0.10, NS not significant (Kruskal-Wallis test followed by Mann-Whitney U test with Bonferroni corrections).

The trends for most demographic variables followed a similar pattern for infection and death clusters except for poverty. For population density and ethnicity (African American or Hispanic American) median values showed a decreasing trend from cluster **1** to cluster **3**, while for age and the distance from epicenter an opposite trend was observed (Figure 3). Furthermore, the difference between clusters **1** and **2** was not significant for these variables but their difference with cluster **3** was found to be significant (except between clusters **2** and **3** for age). Interestingly, for the percentage of the population below the poverty line, the highest median value is observed in cluster **2** for the infection group, and in cluster **1** for the death group. The median values in cluster **3** was intermediate both infection and death groups, suggesting a more complex role of poverty on COVID-19 infection and mortality than other demographic variables.

The clusters in the fatality group demonstrated a different pattern of association with demographic variables. For all variables (except poverty), the median values of cluster **2** were more distant than both clusters **1** and **2**, with the values being significantly higher for population density and ethnicity, and significantly lower for age and the distance from epicenter (Figure 3). While most of these variables’ median values were in proximity for clusters **1** and **2**, percentage of population with age over 55 yr was significantly higher in cluster **1** than cluster **2** (36.55 vs. 33.71), conforming with the existing literature that older age contributes to higher disease fatality. Overall, this analysis suggests the role of demographic structure towards the extent of observed infection, death, and fatality from COVID-19.

### 3.3 Impact of environmental factors on COVID-19

Several recent studies indicated environmental factors such as air pollution and temperature can play a role on COVID-19 transmission and severity (Li et al., 2020; Wu et al., 2020). Therefore, we investigated if temperature and air pollution have any association to the differences observed in COVID-19 landscape across the NYS counties. Temperature and AQI data were collected for the years 2015-2019 from one EPA site representative for each cluster. To better understand the differences of these variables between the clusters, ARIMA models were constructed from time series data. Figure 4 shows ARIMA models of predicted values with 95% confidence bands for temperature and AQI, with the plots comparing models for clusters based on infection, death, or fatality. The low and comparable AIC values for all conditions (Table S1 and S2) confirm the model robustness. Although the model predicted temperature for cluster **3** was lower than other two clusters, considerable overlap of the confidence bands prevented inferring an association with temperature (Figure 4A). Similarly, the confidence bands of the models for temperature in fatality clusters also demonstrated a substantial overlap. The AQI models showed a larger separation of cluster **3** from clusters **1** and **2** for infection and death groups with predicted AQI values for cluster **3** was substantially lower than that of the other two clusters. The AQI values in the clusters for fatality group also demonstrated a broad separation with values for cluster **1** was found to be intermediate between clusters **2** and **3**. Thus, the analyses of data from EPA suggest COVID-19 in NYS is linked to poor air quality but not with outdoor temperature.

**Figure 4.**
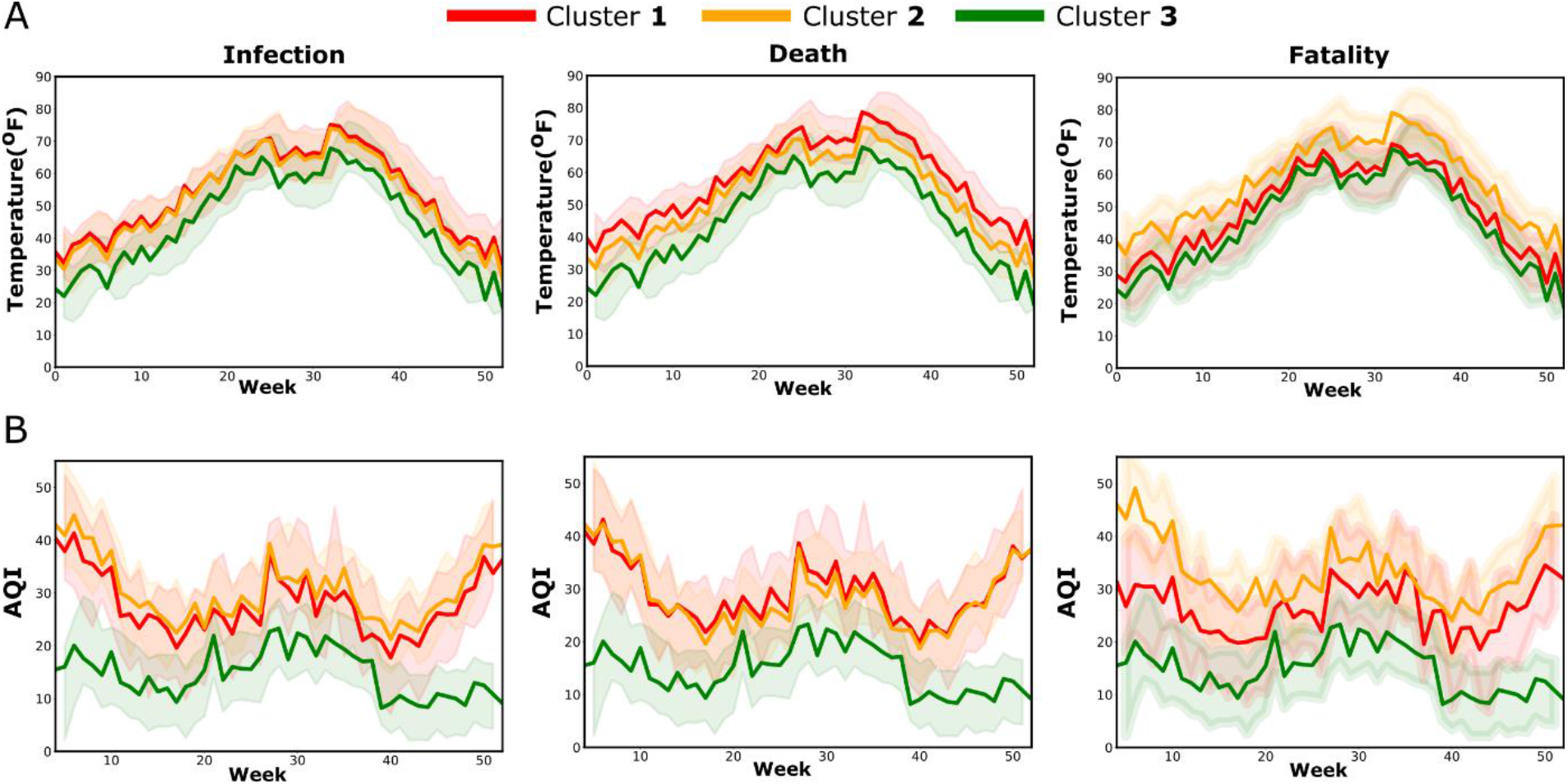
ARIMA time series analysis of temperature (A) and air AQI (B) from weekly EPA data (2015-2019). Predicted values with 95% confidence band for one year, starting from January are shown in the plots. EPA sites for the clusters were located in representative counties for each cluster. Abbreviations: ARIMA, autoregressive integrated moving average; AQI, air quality index.

**Figure 5.**
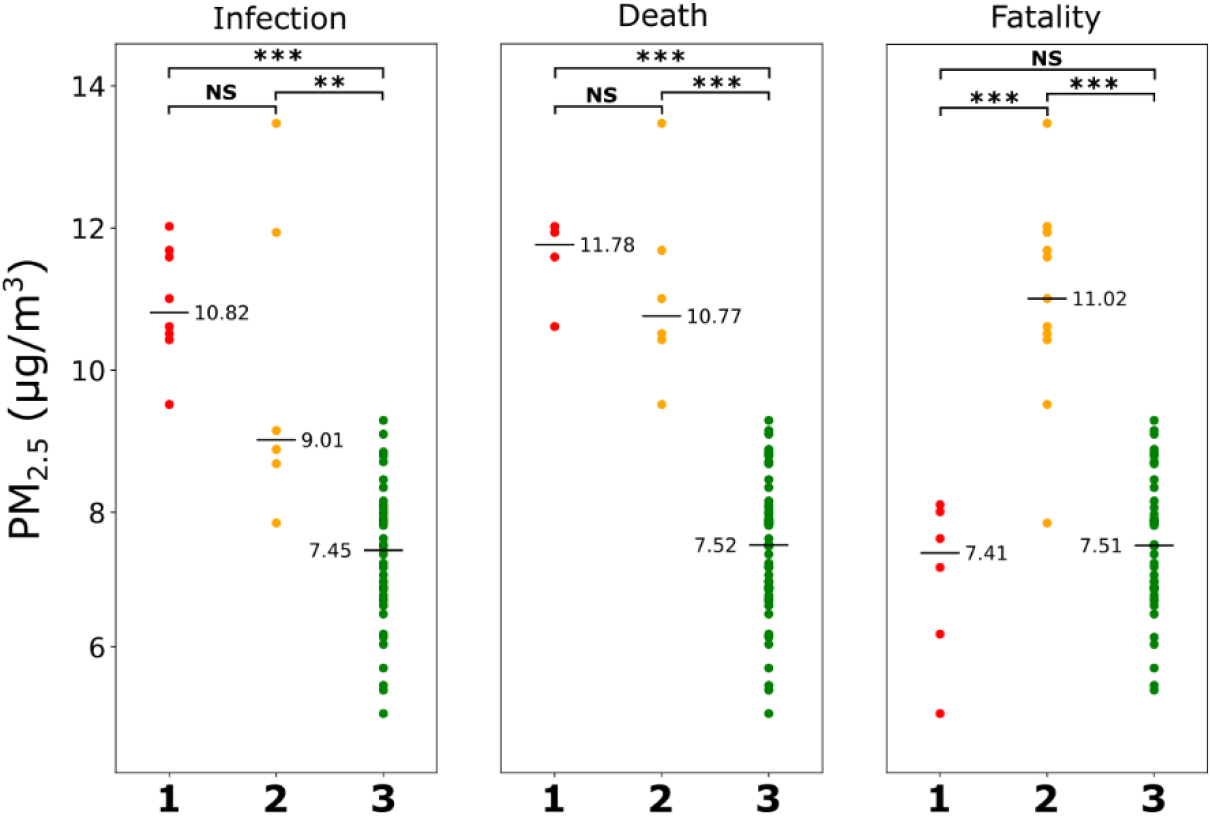
Average PM_2.5_ values of NYS counties estimated from satellite data (2000-2016) are compared between the clusters based on COVID-19 infection, death, and fatality rates. Horizontal lines represent median values. ***P < 0.01, **P < 0.05, *P < 0.10, NS not significant (Mann-Whitney U test with Bonferroni corrections).

EPA sites provide accurate measurement of air quality, however, there are relatively few sampling sites across NYS. To understand the variability of air quality among counties, we used PM_2.5_ values estimated from satellite data over a time period of 2000-2016 (Wu et al., 2020). The findings from satellite data overall corroborate well with the observations from EPA data. PM_2.5_ of cluster **3** was significantly lower than both clusters **1** and **2** for COVID-19 infection and death, with no significant difference between the latter two. For the fatality group, the PM_2.5_ of cluster 1 (high fatality, low infection) was similar to cluster **3** (low fatality, low infection), both being significantly lower than cluster **2**, which includes counties with high infection but lower fatality rate.

### 3.4 Relative contributions of risk factors on COVID-19 infection, death, and fatality

Since clustering of counties based on COVID-19 infection, death, or fatality demonstrated a distinct pattern of association with demographic or environmental risk factors, we wanted to further elucidate the role of these variables on the specific aspects of COVID-19 burden. Six risk factors namely, age above 55 yr, ethnicity (African American and Hispanic American population), poverty, population density, distance from the epicenter, and PM_2.5_ were considered as predictor variables and multivariate regression model with forward “stepwise” selection was used for analysis. Three separate models were constructed using infection, death, and fatality rate as dependent variables to understand the relative contribution of the risk factors for each of these outcomes. Rockland county was excluded from the models as it was identified as an outlier while performing residual analyses of the regression output.

Multicollinearity among the predictor variables was tested before their incorporation in the regression model. Ethnicity and PM_2.5_ were found to have strong positive correlation (r = 0.81, P < 0.001, Figure 6A). Further, VIF calculated for risk variables showed ethnicity to have a value higher than the acceptable cut-off value of 5 for infection and death rates, and thus not included in the final models. The models revealed distinct contributions of the risk factors to infection, death, and fatality rates (Figure 6B). PM_2.5_ and distance from the epicenter were found to be the two most important predictors for infection and death. For infection rate, distance from the epicenter was the strongest predictor with an adjusted R^2^ value of the model was 0.60 when considered as a sole contributor, with the value increased to 0.71 upon adding PM_2.5_ as a predictor variable (Figure 6B,C). Similarly, PM_2.5_ was the strongest predictor for death rate with an adjusted R^2^ value of 0.69 considered alone and the value increased to 0.73 following the inclusion of distance from the epicenter. Adding population density, age, and poverty to the regression model only minimally increased the adjusted R^2^ value, with the final values being 0.74 and 0.73 for infection and death rates, respectively.

**Figure 6.**
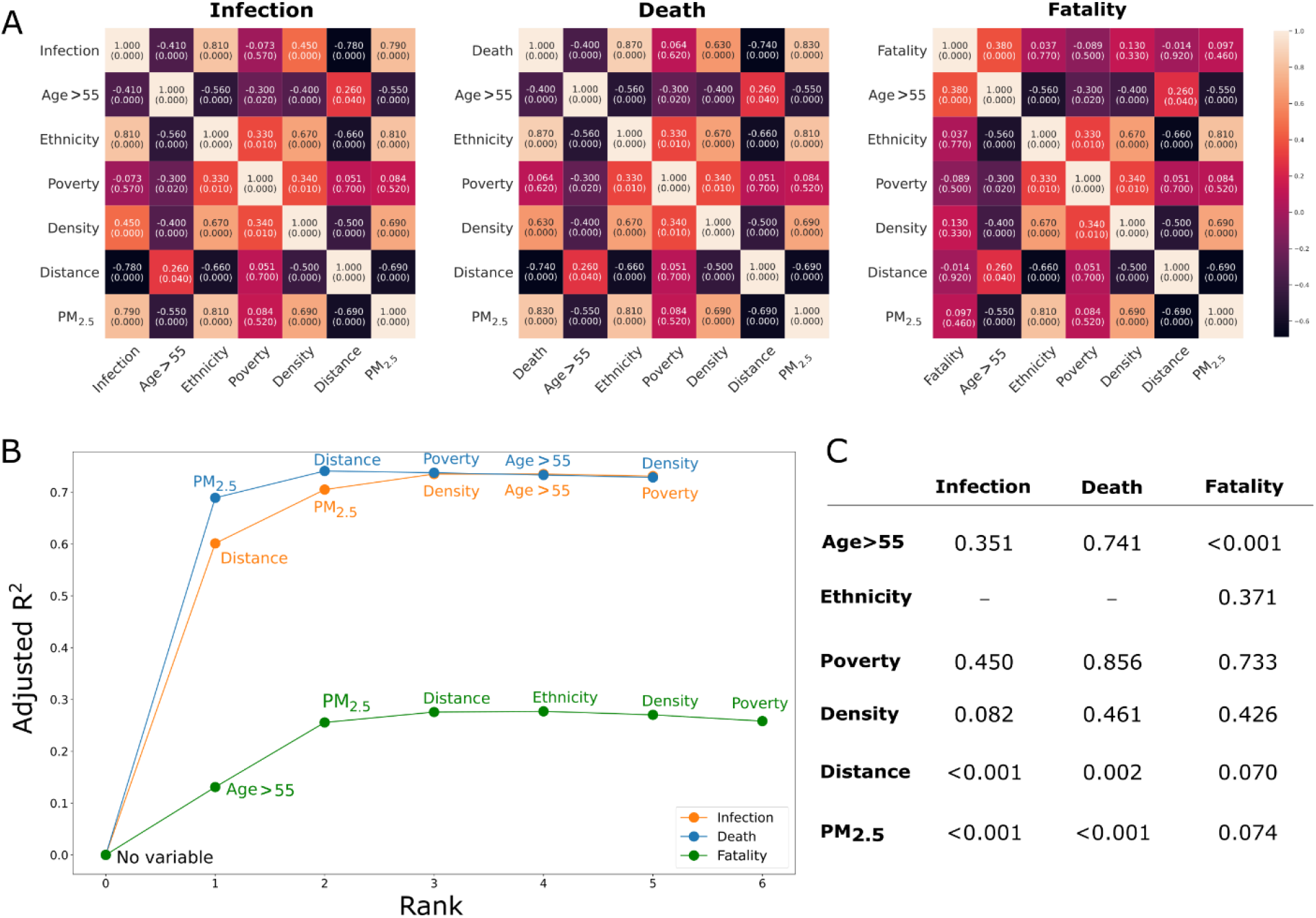
Regression analysis to assess relative contribution of risk factors on COVID-19. (A) Correlation matrices between demographic risk variables and PM_2.5_ for infection, death, and fatality. Pearson’s correlations coefficients between the variables are shown with corresponding P values are mentioned in the parentheses. (B) Stepwise regression model with forward selection after removal of the risk factors having strong multicollinearity. (C) Table showing the P values of the regression coefficients from the analysis. Abbreviations used: Density, population density (per mi^2^); Ethnicity, African American and Hispanic American (%); Poverty, population below poverty line (%); Distance, distance from the epicenter (mi).

In contrast to infection and death rates, in the regression model for fatality rate, age over 55 was found to be the strongest predictor, followed by PM_2.5_, and distance from the epicenter (Figure 6C). However, the adjusted R^2^ value for the final model was 0.26, suggesting the goodness of model fit for fatality rate is much lower than for infection and death rate. Together, the regression analysis helped to understand the relative impact of various risk factors on different aspects of COVID-19 burden.

## 4 Discussion

Our analysis has shown a wide heterogeneity in the infection, death, and fatality rates from COVID-19 among the counties in the NYS during the first pandemic wave. Infection was found to be strongly correlated with death but not with fatality. We observed that infection, death, and fatality rates have a significant association with air quality and various demographic factors.

Furthermore, regression analysis study has revealed a differential contribution of these factors on the infection, death, and fatality rates. These results could help to better understand the impact of environmental and demographic factors on COVID-19 in NYS.

An association of PM_2.5_ with infection, death, and fatality rates is observed in our analysis. This finding is in agreement with studies focused on the role of outdoor air pollution on COVID-19 transmission and health outcomes (Benmarhnia, 2020; Gupta et al., 2020; Lolli et al., 2020; Pozzer et al., 2020; Wu et al., 2020). PM_2.5_ has been reported to be positively correlated with both increased COVID-19 transmission and mortality rates. The effect on mortality is attributed to chronic exposure to higher PM_2.5_, which can lead to multiple respiratory and cardiovascular illnesses (Benmarhnia, 2020; Pozzer et al., 2020; Wu et al., 2020). While increased virus transmission can be partly attributed to the higher susceptibility to infection due to the presence of comorbid conditions, acute lung inflammation because of air pollution could also play a role. It is to be noted that the average air quality across NYS is good, with the average PM_2.5_ values meeting the safe limit (< 12 µg/m^3^) set by EPA, the association with adverse effects of COVID-19 can be clearly discerned. Although outdoor air temperature is also reported to have an association with COVID-19 transmission (Lolli et al., 2020), such association was not evident in NYS data.

Interestingly, we observed a strong correlation between PM_2.5_ and the percentage of the population belonging to African American or Hispanic American ethnicity, indicating they are exposed to a higher level of PM_2.5_ than the average population. Multiple studies have reported that these two ethnicities in the US are at disproportionately higher risk to be affected by COVID-19 (Cordes and Castro, 2020; Li et al., 2020; Martinez et al., 2020; Yancy, 2020). Factors related to socioeconomic inequities such as the greater risk of virus exposure from professional demand or living in crowded accommodation, higher prevalence of chronic comorbidity, and restricted access to healthcare are thought to underlie such differences (Patel et al., 2020). Our results suggest that exposure to air pollution could be a contributor to further increase this disparity. Interestingly, low socioeconomic status is known to pose a greater risk for COVID-19 exposure; however, for NYS, we observed counties in cluster **3** for infection and death rates to have a higher proportion of people living below the poverty line than the other two counties. That cluster **3** counties have a relatively lower risk from other demographic and environmental variables could explain this apparent discrepancy. Indeed, when considering the population of NYC alone, poverty and COVID-19 are found to be positively correlated (Cordes and Castro, 2020).

Two demographic variables—distance from the epicenter and age above 55 years—have demonstrated substantial relative contribution in our regression models. However, their influences are distinct. The distance of counties from the disease epicenter is inversely associated with COVID-19 infection and death rate. This finding is not unusual as the disease spread would be facilitated by the population mobility with the highest effect on the neighboring regions. On the other hand, in the regression model of fatality, age over 55 years plays is found to be the most significant independent variable. Fatality from COVID-19 depends on the health of patients where age plays a crucial role. Increased risk of the aged population to complications and death from COVID-19 is observed across the world, and multiple factors including the existence of chronic comorbid conditions and a weaker immune system are thought to underlie such vulnerability.

Grouping the counties into clusters not only helped to visualize how NYS counties are impacted by specific COVID-19 adversities but also allowed easier comparison of their association with various demographic and environmental risk variables. While the regression models have further helped to understand the relative contribution of these risk variables on the disease, it should also be noted that the adjusted R^2^ value of the regression model for fatality rate is substantially lower than the models for infection and death rate (0.26 *vs*. 0.74 and 0.73). This difference suggests the need to include additional variables in our model that might be critical in determining COVID-19 fatality. Such variables could be measures pertaining to the outcome of an infected individual, including the availability and access to healthcare resources for advanced COVID-19 management and public awareness for early diagnosis and treatment.

## Conclusions

We have analyzed here the impact of contextual factors on COVID-19 in NYS during the first pandemic wave. Air pollution and multiple demographic factors are found to have a conspicuous but distinct association with the variability of COVID-19 burden observed across the counties. Exposure to a higher level of PM_2.5_ demonstrated association with all COVID-19 estimates considered, while the distance from the epicenter and increased age are predominantly related to disease transmission and severity of health outcome, respectively. The regression model helped to estimate the relative contribution of these factors in infection. Understanding the interplay of these risk variables would help in better assessment of the disease impact in the population and in developing preventative or mitigative measures to contain infection.

## Data Availability

Raw datasets are publicly available for download. The details of the sources and links to the sites are provided in a table in the manuscript.

## CRediT authorship contribution statement

**Sumona Mondal**: Writing – original draft, Supervision, Methodology, Formal analysis, Project administration.

**Chaya Chaipitakporn**: Formal analysis, Visualization, Data Curation.

**Bridget Wangler**: Visualization.

**Vijay Kumar**: Data curation.

**Supraja Gurajala**: Review and editing.

**Suresh Dhaniyala**: Validation, Review and editing.

**Shantanu Sur**: Conceptualization, Investigation, Supervision, Writing -review and editing.

## Declaration of conflicts of interest

The authors declared that they have no conflict of interests.

### Acknowledgments

Vijay Kumar acknowledges the support from US-Pakistan Knowledge Corridor PhD Scholarship Program under Higher Education Commission, Pakistan. Bridget Wangler thanks the Clarkson University Honors Program for their support.

